# Serial measurements of Growth Differentiation Factor-15 in patients with Non-ST-Elevation Acute Coronary Syndrome and recurrent ischemic events

**DOI:** 10.64898/2026.10.11.26364375

**Authors:** Ke Chen, Runan Zhang, Xinyi Rao, Junyi Wu, Hui Yuan

## Abstract

**Background:** Growth differentiation factor 15 (GDF-15) has emerged as a prognostic biomarker in patients with non-ST-segment-elevation acute coronary syndrome (NSTE-ACS). Our study evaluated the temporal evolution of GDF-15 in the acute phase and its association with recurrent ischemic events.

**Methods:** Serum samples were collected from patients with NSTE-ACS at admission and at 1 and 3 hours after admission. GDF-15 levels, along with other prognostic biomarkers, NT-proBNP and hs-cTnT, were measured. Spearman’s rank-correlation coefficients and linear regression models were used to identify the variables associated with GDF-15 levels at admission and with their temporal changes. Associations between GDF-15 and clinical outcomes were evaluated using Cox proportional hazards models. To assess incremental prognostic value, the areas under the curve (AUC) were calculated for each model.

**Results:** GDF-15 levels showed a slight increasing trend within 3 hours of admission. The 1-hour change in GDF-15 was correlated with hs-cTnT changes. Increased 1-hour GDF-15 levels, compared with the admission levels, were associated with a higher risk of recurrent ischemic events during the 1-year follow-up (1.9% vs. 17.5%, *P* =0.019) and were an independent predictor after adjusting for established clinical variables and biomarkers (adjusted hazard ratio, 1.71 [1.28 to 228.75], *P* =0.03). Furthermore, serial measurements of GDF-15 provided additional prognostic information beyond traditional risk factors.

**Conclusions:** Serial measurements of GDF-15 of NSTE-ACS patients provided independent and incremental prognostic value for predicting recurrent ischemic events. These findings support the potential utility of GDF-15 as a dynamic biomarker for early risk stratification in the acute management of NSTE-ACS.

## 1 Introduction

The risk of recurrent ischemic events in patients with non-ST-elevation acute coronary syndrome (NSTE-ACS) patients has been significantly decreased due to advances in routine reperfusion therapy and optimized secondary preventive treatments; However, a considerable proportion of patients remain at high risk for recurrent ischemic events and cardiovascular death [1]. The risk of adverse events varies considerably across the spectrum of the patients with NSTE-ACS. Early and continuous risk stratification, incorporating clinical characteristics and biomarkers, is essential for informed decision-making and improving the prognosis of patients with NSTE-ACS.

Growth differentiation factor-15 (GDF-15), a 25 kDa secreted homodimeric protein, is a stress-responsive member of the transforming growth factor β cytokine superfamily and has been identified as a pleiotropic protein involved in embryonic development, inflammatory regulation, cellular stress responses, and tissue repair following acute injury [2, 3]. Experimental ischemia and reperfusion injury and pressure overload have been found to stimulate the expression and secretion of GDF-15 in cultured cardiomyocytes and mouse heart [4, 5]. In a retrospective analysis of a randomized controlled trial, GDF-15 has shown independent prognostic value in predicting death in patients with NSTE-ACS. Additionally, in this study, the tertiles of GDF-15 levels were used as cutoff points to identify patient subgroups at varying risk levels for death: low risk (< 1200 ng/L), intermediate risk (1200 to 1800 ng/L) and high risk (>1800 ng/L) [6]. Furthermore, the prognostic role of GDF-15 in patients with cardiovascular disease has garnered widespread attention [7–11].

Previous studies have primarily focused on the prognostic value of single-measurement GDF-15 levels across different conditions of acute coronary syndrome (ACS) condition [12–14]. Although studies have shown that GDF-15 changes in patients with NSTE-ACS after disease stabilization have no correlation with prognosis [13], the risk stratification of GDF-15 may be enhanced by considering dynamic changes in its levels over time during the acute phase. Therefore, this analysis aimed to assess the time course of GDF-15 levels and the association between changes in GDF-15 levels and the onset of subsequent recurrent ischemic events during the acute phase of NSTE-ACS.

## 2 Materials and Methods

### 2.1 Study Population

This retrospective cohort study enrolled patients presenting with non-traumatic chest pain or other symptoms suggestive of acute myocardial infarction (MI), who were admitted to the emergency department of Beijing Anzhen Hospital, Capital Medical University, between 01/05/2017 and 31/05/2020. The data were accessed for research purposes between 25/08/2022 and 13/09/2022. Eligible patients were defined as those diagnosed with NSTE-ACS according to the 2015 ESC Guidelines for managing of acute coronary syndromes in patients presenting without persistent ST-segment elevation [15]. Diagnosis was established based on clinical presentation and physical examination, combined with electrocardiogram and high-sensitivity cardiac troponin; it was eventually confirmed via coronary angiography. Serum samples were available from patients on admission and at 1- and 3-hours post-admission. Exclusion criteria included electrocardiographic evidence of ST-segment elevation, pregnancy, and severe hepatic or renal dysfunction. During the 1-year follow-up, recurrent ischemic events were documented, including MI and unstable angina leading to hospitalization, which were evaluated by patient records and telephone contacts. During data processing, all records were anonymized by removing directly identifiable information.

### 2.2 Laboratory Analysis

Venous blood samples were collected on admission, after 1 hour, and after 3 hours. The samples were centrifuged, and serum was stored frozen at -80°C for further analyses. ElectroChemi-Luminescence Immunoassays were used to measure GDF-15 (Elecsys GDF-15 assay, Roche Diagnostics International Ltd., Rotkreuz, Switzerland) on cobas e801 analytical unit, with intra-assay imprecision ranging from 0.8% to 1.5% and within-laboratory imprecision ranging from 4.0% to 12.2% for samples containing 670 to 16039 ng/L [16]. N-terminal pro B-type natriuretic peptide (NT-proBNP) (Roche Diagnostics, Elecsys assay) and hypersensitive cardiac troponin T (hs-cTnT) (Roche Diagnostics, Elecsys assay) were analyzed with the ElectroChemi-Luminescence Immunoassays. Hypersensitive C reactive protein (hs-CRP) (Cardiac C-Reactive Protein (Latex) High Sensitive, Roche Diagnostics GmbH, Mannheim, Germany) and serum creatinine (Cr) (Creatinine Jaffé Gen.2, Roche Diagnostics GmbH, Mannheim, Germany) were obtained from the laboratory information system (LIS); However, and only admission levels were available. All biomarker measurements were conducted by laboratory personnel who were blinded to clinical diagnosis and outcomes.

### 2.3 Statistical Analysis

All statistical analyses were conducted using R software for macOS (Version 4.2.1, https://www.r-project.org/). Continuous variables were described using the median and interquartile range (25th–75th quartile) and were compared using either a t test or the Mann-Whitney. Categorical variables were presented as frequency and percentage and compared using the χ² test. Spearman’s rank correlation coefficients were calculated to evaluate the relationships between GDF-15 levels on admission and baseline characteristics, as well as the concentrations of other biomarkers. Additionally, the correlations between the changes in GDF-15 levels and the changes in NT-proBNP and hs-cTnT levels were assessed. Multiple linear regression analysis was performed to evaluate the association between admission GDF-15 levels and other predictors. Variables that were not normally distributed (GDF-15, NT-proBNP, hs-cTnT, hs-CRP, and Cr) were transformed to their natural logarithms prior to conducting multiple linear regression analyses. The Wilcoxon signed rank test were used to compare the temporal evolution of GDF-15 within the patient group. Differences in the proportions of outcome events across different strata of GDF-15 levels and changes in GDF-15 levels were assessed using Fisher’s exact test. Restricted cubic splines with 4 knots were used to determine appropriate cutoff values for changes in GDF-15 levels. The Kaplan-Meier method was employed to illustrate the timing of events during follow-up based on the temporal evolution of GDF-15 levels. Statistical comparisons of event timing between groups were conducted using the log-rank test. Simple Cox regression analysis was used to identify predictors of recurrent ischemic events during the 1-year follow-up period. Subsequently, all clinical characteristics were included in multiple Cox regression analyses to assess the independent associations with recurrent ischemic events. Measurements of receiver operating characteristic curves were conducted to explore the additional prognostic value of the change in GDF-15 levels compared to GDF-15 levels on admission. The DeLong test was used to compare the differences in the areas under the curve (AUC) between the models. All P-values were two-sided, and a value of <0.05 was considered statistically significant.

A post hoc power analysis was performed using PASS 2021 based on the hazard ratio from the univariate Cox model. With 92 patients (40 in the ΔGDF-15 > 0 group and 52 in the Δ ≤ 0 group) and an observed HR of 9.71, the estimated power was 88.97% at a two-sided alpha level of 0.05. This suggests the study had sufficient power to detect the observed survival difference.

Informed consent was obtained from all patients or waived as appropriate, and the study protocol was approved by the Ethics Committee of Beijing Anzhen Hospital, Capital Medical University (2021-072). For participants whose data could not be retrieved from electronic medical records, verbal consent was obtained via telephone, documented in writing and witnessed according to institutional guidelines. For participants whose anonymized data were available through the hospital system, the requirement for consent was waived by the Ethics Committee.

## 3 Results

### 3.1 Baseline Characteristics

This prospective cohort consisted of 68 men (73.91%) and 24 women (26.09%) with a median age of 64 (58-70) years. The clinical characteristics and laboratory data of this patient sample are summarized in Table 1. About two-thirds of the patients were diagnosed with unstable angina, and more than half had comorbidities of hypertension and hyperlipidemia.

**Table 1.** Baseline Characteristics of Patients in Relation to the Temporal Evolution of GDF-15.

| Characteristics | Combined (N=92) | Temporal Evolution of GDF-15 <sup>a</sup> |  | P Value |
| --- | --- | --- | --- | --- |
| | | $\Delta > 0$ (N=40) | $\Delta \leq 0$ (N=52) | |
| Age, y | 64 (58-70) | 64 (58-70) | 66 (57-70) | 0.972 |
| Gender, %(n) |  |  |  | 0.610 |
| Male | 68 (73.91%) | 28 (70.00%) | 40 (76.92%) |  |
| Female | 24 (26.09%) | 12 (30.00%) | 12 (23.08%) |  |
| BMI, kg/m<sup>2</sup> | 26.42 (24.16-28.15) | 25.40 (24.00-28.70) | 26.83 (24.70-28.04) | 0.464 |
| DBP, mmHg | 75.0 (68.0-82.0) | 76.5 (67.0-81.5) | 74.5 (68.0-82.0) | 0.850 |
| SBP, mmHg | 129.5 (120.0-140.0) | 129.0 (120.0-137.0) | 129.5 (120.0-140.0) | 0.570 |
| Smoke, %(n) |  |  |  | 0.722 |
| Never | 35 (38.04%) | 16 (40.00%) | 19 (36.54%) |  |
| Past | 27 (29.35%) | 10 (25.00%) | 17 (32.69%) |  |
| Current | 30 (32.61%) | 14 (35.00%) | 16 (30.77%) |  |
| Diagnosis of NSTEMI-ACS, % (n) |  |  |  | 0.567 |
| UA | 65 (70.65%) | 30 (75.00%) | 35 (67.31%) |  |
| NSTEMI | 27 (29.35%) | 10 (25.00%) | 17 (32.69%) |  |
| Treatment strategies, % (n) |  |  |  | 0.0735 |
| Invasive strategy | 42 (45.65%) | 23 (57.50%) | 33 (63.46%) |  |
| Conservative strategy | 50 (54.35%) | 17 (42.50%) | 19 (36.54%) |  |
| Medical history, % (n) |  |  |  |  |
| **Hypertension** | 70 (76.09%) | 31 (77.50%) | 39 (75.00%) | 0.974 |
| **Diabetes mellitus** | 38 (41.30%) | 21 (52.50%) | 17 (32.69%) | 0.093 |
| **Hyperlipidemia** | 88 (95.65%) | 37 (92.50%) | 51 (98.08%) | 0.433 |
| **Myocardial infarction** | 16 (17.39%) | 5 (12.50%) | 11 (21.15%) | 0.419 |
| **Previous PCI** | 34 (36.96%) | 14 (35.00%) | 20 (38.46%) | 0.902 |
| **Chronic kidney disease** | 4 (4.35%) | 2 (5.00%) | 2 (3.85%) | 1.000 |
| **Laboratory analyses** |  |  |  |  |
| **NT-proBNP, ng/L** | 96.55<br>(49.88-228.75) | 103.25<br>(51.10-197.75) | 92.35<br>(48.03-228.75) | 0.440 |
| **hs-cTnT, ng/L** | 10.15 (6.15-27.53) | 11.60 (5.43-27.95) | 9.72 (6.49-25.50) | 0.838 |
| **hs-CRP, mg/L** | 1.35 (0.84-3.10) | 1.56 (0.84-3.10) | 1.33(0.88-2.83) | 0.856 |
| **Cr, μmol/L** | 77.60 (67.55-89.95) | 81.35 (67.95-90.08) | 76.45 (66.53-89.73) | 0.625 |

<sup>a</sup> Temporal evolution of GDF-15 from admission to 1 hour after admission.
Abbreviations: BMI, body mass index; DBP, diastolic blood pressure; SBP, systolic blood pressure;
NSTE-ACS, non-ST-segment elevation acute coronary syndrome; UA, unstable angina; NSTEMI,
non-ST-segment elevation myocardial infarction; PCI, percutaneous coronary intervention;
NT-proBNP, N-terminal pro-B-type natriuretic peptide; hs-cTnT, high-sensitivity cardiac troponin
T; hs-CRP, high-sensitivity C-reactive protein; Cr, creatinine; GDF-15, growth differentiation factor

### 3.2 Temporal Evolution of GDF-15 Levels in the Acute Phase of NSTE-ACS

GDF-15 levels on admission ranged from 400 to 9698 ng/L with a median of 1294 (858-1593) ng/L. A total of 39 (42.4%), 38 (41.3%), and 15 (16.3%) of the patients presented with GDF-15 levels <1200 ng/L, between 1200 and 1800 ng/L, and >1800 ng/L, respectively. Although GDF-15 levels on admission had a strong correlation with GDF-15 levels at later time points, there was a slight increasing trend at 3 hours (*P* =0.086) (Table 2).

**Table 2.**
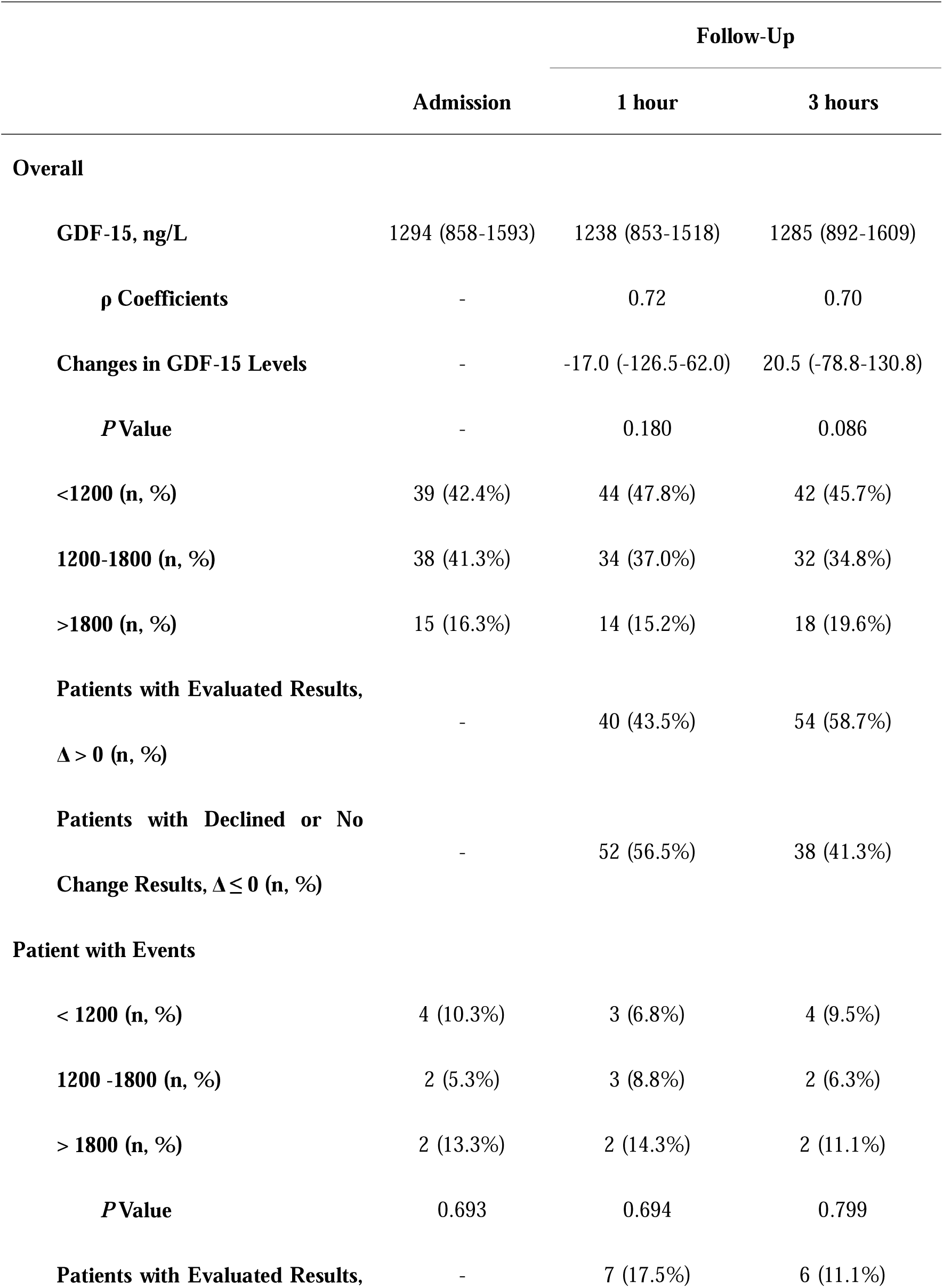
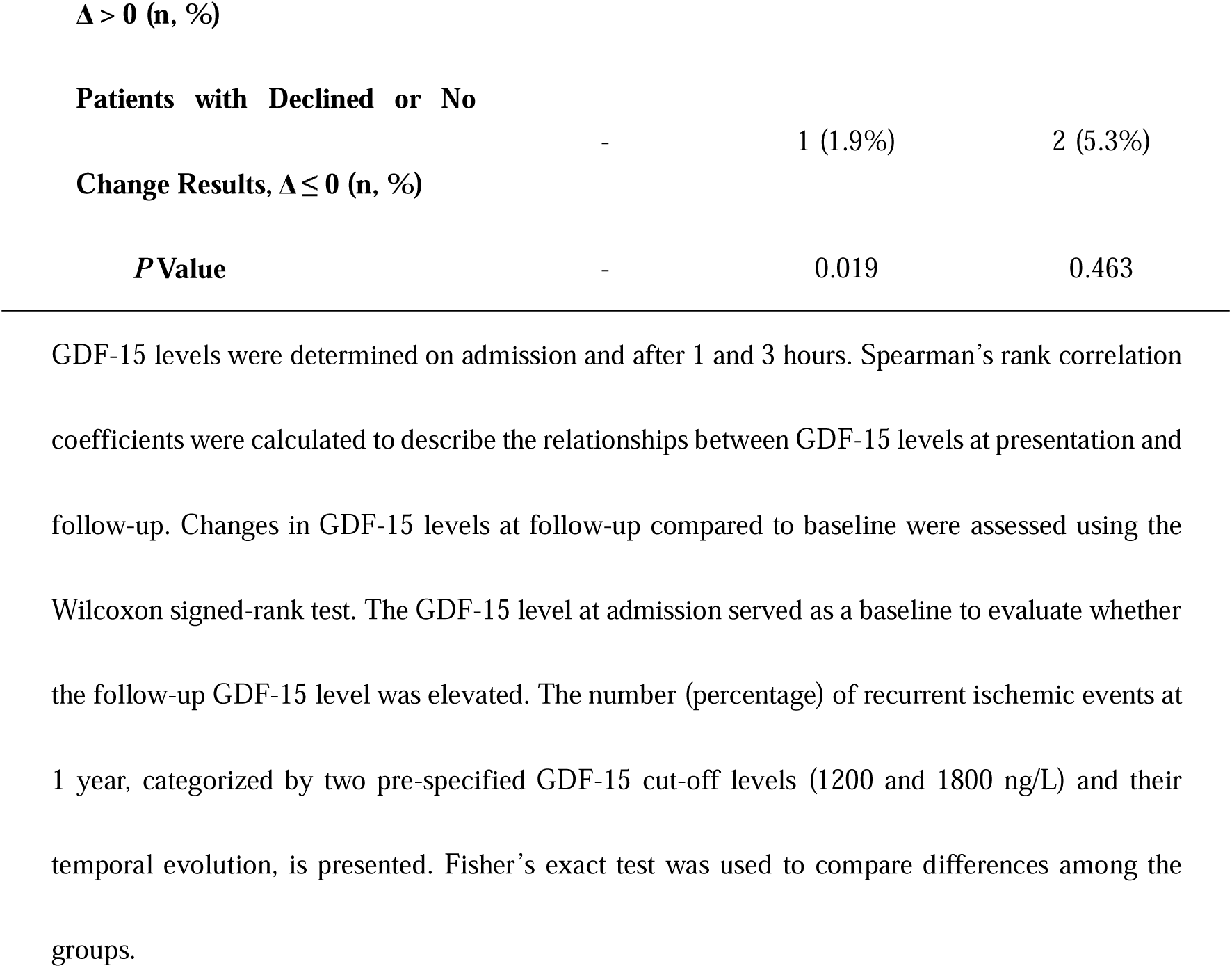
Temporal Evolution of GDF-15 Levels in the Acute Setting of NSTE-ACS and 1-year Primary Events.

The restricted cubic spline analysis revealed a nonlinear relationship between the 1-hour change in GDF-15 levels and the risk of recurrent ischemia (*P* < 0.001). When the change in GDF-15 levels was close to zero, the hazard ratio for outcomes was greater than 1 (Fig 1). Based on this finding, zero was selected as the threshold for the 1-hour change in GDF-15 levels. The continuous variable was subsequently converted into a categorical variable for further analysis. Although the 3-hour change in GDF-15 levels showed a linear relationship with the hazard ratio (not shown), for consistency with the 1-hour analysis, the 3-hour GDF-15 change value was also converted into a categorical variable using a threshold of zero. At 1 hour and 3 hours, 43.5% and 58.7% of patients had elevated GDF-15 levels compared with baseline, respectively (Table 2).

**Fig 1.**
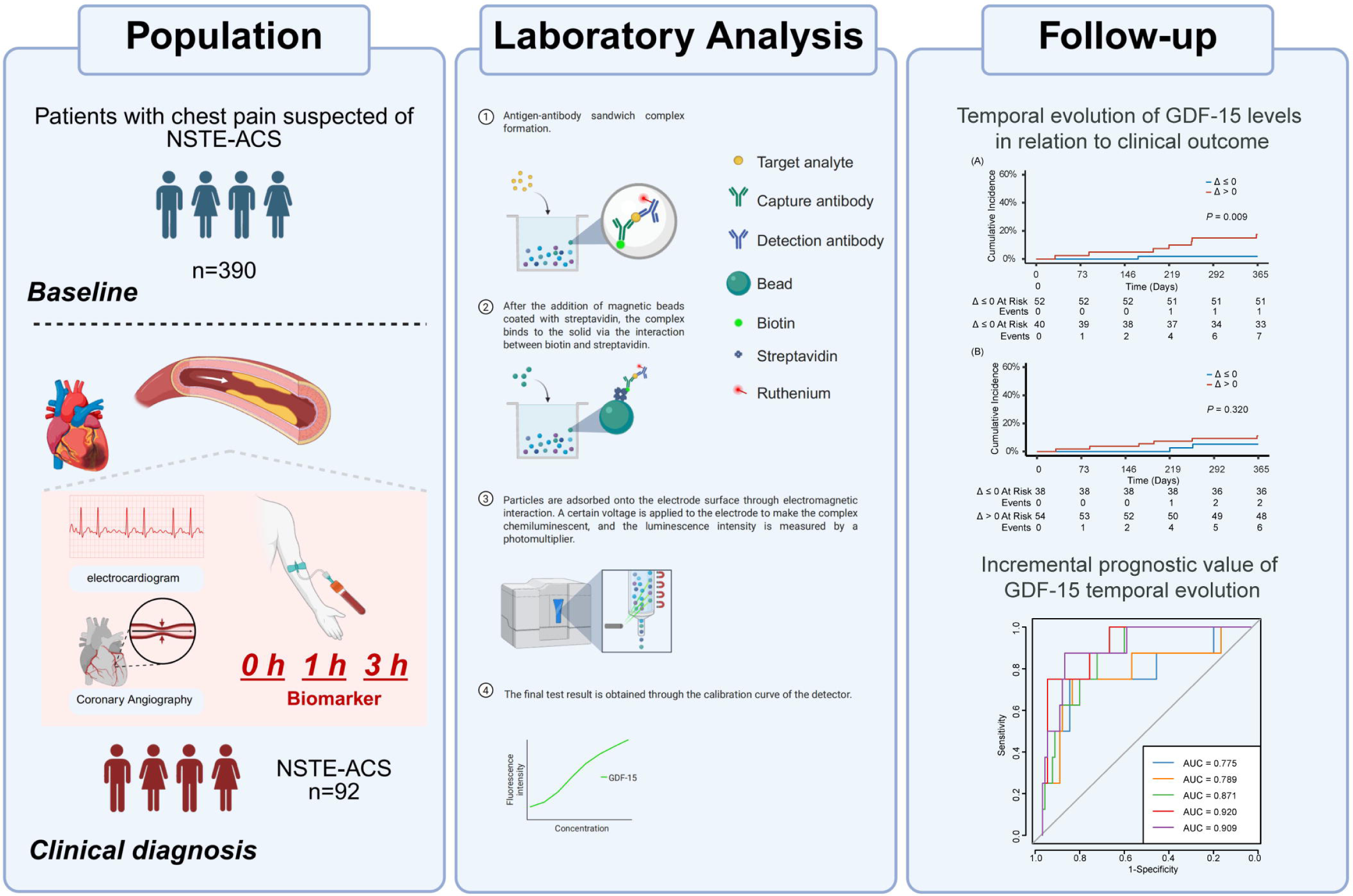
Restricted cubic splines analysis showing the association between 1-hour change in GDF-15 levels and HR for clinical outcome. The X-axis represents the absolute change in GDF-15 levels (pg/mL) within one hour, while the Y-axis shows the estimated HR and its 95% CI for adverse outcomes. The horizontal dashed line at HR = 1 indicates no effect. The vertical dashed line is drawn at the intersection where the red curve crosses the HR = 1 line, indicating the threshold value of one-hour GDF-15 increase associated with a significantly elevated risk. This threshold suggests that when GDF-15 levels increase beyond this point within one hour, the risk of adverse outcomes rises notably. Abbreviations: HR, hazard ratio; CI, confidence interval.

### 3.3 GDF-15 Levels and Temporal Evolution in Relation to Clinical Variables and Biomarkers

GDF-15 levels on admission were positively associated with age, NT-proBNP levels on admission, and Cr, with no relationship to the levels of hs-cTnT on admission (Table 3). Multiple linear regression analysis demonstrated that NT-proBNP levels on admission (*P* = 0.006) and Cr (*P* = 0.028) were independently associated with GDF-15 levels. The R^2^ value of this model was 0.31. The correlation coefficients between the change in GDF-15 and NT-proBNP levels at 1 hour and 3 hours were 0.242 (*P* = 0.020) and 0.319 (*P* = 0.002), respectively. Additionally, there was a positive relationship between the change in GDF-15 and hs-cTnT at 1 hour, while no significant relationship was observed at 3 hours (Table 3).

**Table 3.** Relation Between Change of GDF-15 Levels and Clinical and Biochemical Factors in NSTE-ACS.

|  | Spearman's Correlation |  | Multiple Linear Regression |  |
| --- | --- | --- | --- | --- |
| | $\rho$ | P Value | $\beta$ | P Value |
| **GDF-15, ng/L (Baseline)** |  |  |  |  |
| Age, y | 0.210 | 0.045 | 0.004 | 0.605 |
| Male, % | 0.090 | 0.394 | 0.147 | 0.343 |
| BMI, kg/m<sup>2</sup> | -0.052 | 0.625 | -0.001 | 0.456 |
| SBP, mmHg | 0.177 | 0.091 | 0.0002 | 0.963 |
| Current Smoke, % | -0.008 | 0.942 | -0.010 | 0.902 |
| Hypertension, % | 0.171 | 0.104 | 0.174 | 0.178 |
| Diabetes mellitus, % | 0.131 | 0.212 | 0.115 | 0.317 |
| Hyperlipidemia, % | 0.048 | 0.648 | 0.183 | 0.509 |
| Myocardial Infarction, % | 0.092 | 0.381 | -0.082 | 0.594 |
| Chronic Kidney Disease, % | 0.116 | 0.269 | -0.343 | 0.300 |
| Previous PCI | 0.079 | 0.453 | 0.031 | 0.811 |
| NT-proBNP, ng/L (Baseline) | 0.414 | < 0.001 | 0.140 | 0.006 |
| hs-cTnT, ng/L (Baseline) | 0.170 | 0.106 | 0.038 | 0.433 |
| hs-CRP, mg/L | 0.005 | 0.961 | -0.006 | 0.209 |
| Cr, μmol/L | 0.288 | 0.005 | 0.008 | 0.028 |
| **Change Values of GDF-15, ng/L <sup>a</sup>** |  |  |  |  |
| Change Value of NT-proBNP, ng/L | 0.242 | 0.020 | - | - |
| **Change Value of hs-cTnT, ng/L** | 0.337 | 0.001 | - | - |
| **Change Values of GDF-15, ng/L <sup>b</sup>** |  |  |  |  |
| **Change Value of NT-proBNP, ng/L** | 0.319 | 0.002 | - | - |
| **Change Value of hs-cTnT, ng/L** | 0.0850 | 0.421 | - | - |

Spearman's rank correlation coefficients were used to evaluate the association between admission GDF-15 levels and baseline parameters, as well as the 1- and 3-hour GDF-15 changes and other biomarker changes. Multiple linear regression analysis was performed to identify factors independently associated with GDF-15 on admission. <sup>a</sup> The change values of GDF-15 between admission and 1 hour; <sup>b</sup> the change values of GDF-15 between admission and 3 hours.

### 3.4 GDF-15 Levels and Temporal Evolution in Relation to Clinical Outcome

The risk of recurrent ischemic events did not show a significant increase with higher levels of GDF-15 on admission (*P* = 0.693), at 1 hour (*P* = 0.694), or at 3 hours (*P* = 0.799), as shown in Table 2. With the threshold established through restricted cubic spline analysis, 52 patients had a 1-hour GDF-15 change value less than or equal to zero, while 40 patients had a change value greater than zero (Table 1). The baseline characteristics between the two cohorts showed no significant differences. The incidence of recurrent ischemic events was 1.9% (1 of 52) and 17.5% (7 of 40) in the two cohorts (P = 0.019) (Table 2). Among the two cohorts defined by the 3-hour GDF-15 change values, 5.3% and 11.1% of patients, respectively, experienced outcome events (*P* = 0.463). The cumulative probability of outcomes during the 1-year follow-up, stratified by the 1-hour and 3-hour GDF-15 change values, is illustrated in Fig 2.

**Fig 2.**
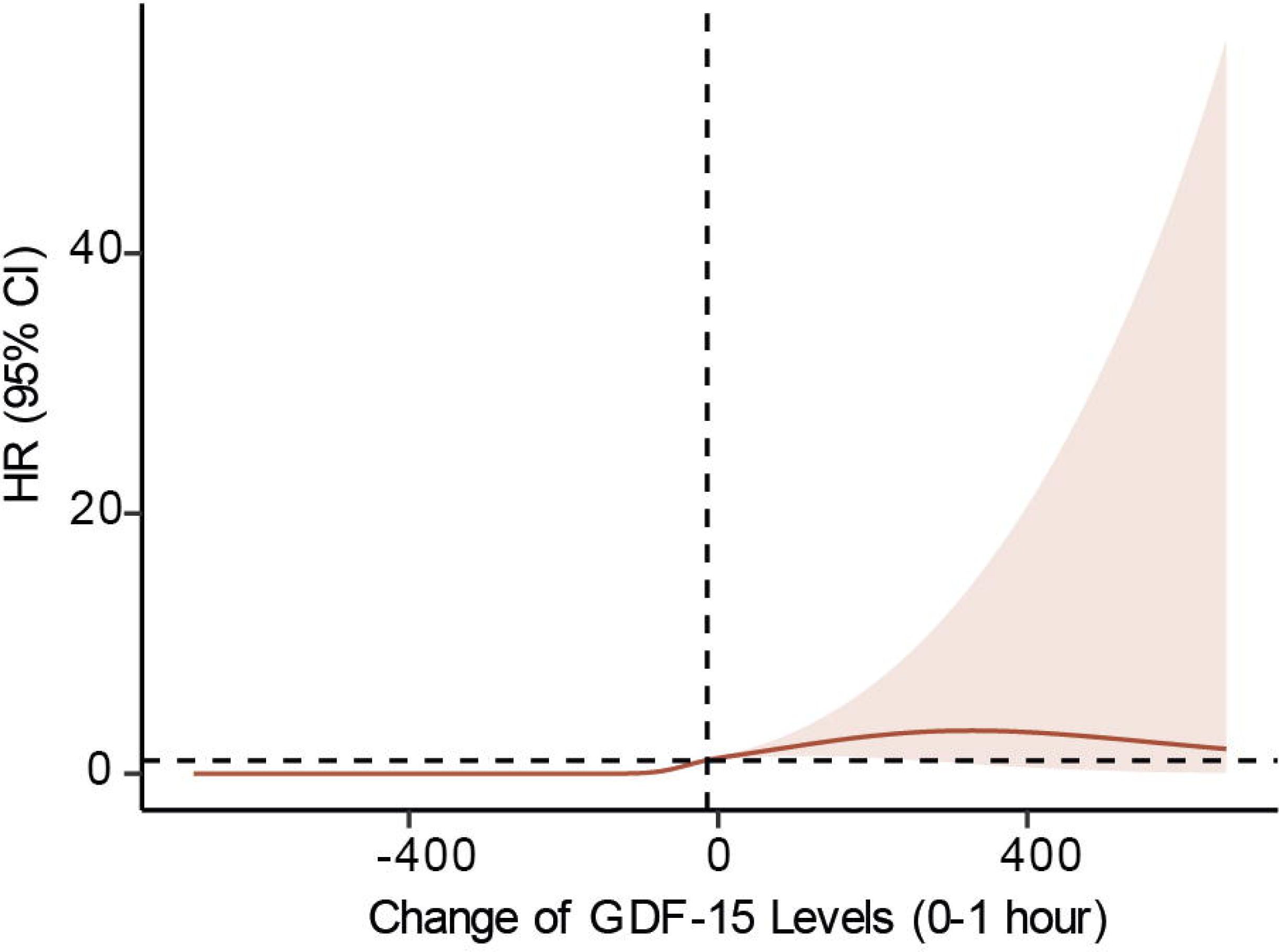
**Kaplan-Meier cumulative incidence curves for recurrent ischemic events during 1-year follow-up**. (A) Cumulative incidence stratified by 1-hour changes in GDF-15 levels (Δ ≤ 0 vs. Δ > 0). Patients with Δ > 0 had a significantly higher risk of recurrent ischemic events compared to those with Δ ≤ 0 (P = 0.009). (B) Cumulative incidence stratified by 3-hour changes in GDF-15 levels (Δ ≤ 0 vs. Δ > 0). No significant difference was observed between the groups (*P* = 0.320). The x-axis represents follow-up time in days, and the y-axis indicates the cumulative incidence (%). The number of patients at risk and the number of events at selected time points are displayed below each curves.

Through simple Cox regression analysis, only 1-hour GDF-15 change values were related to 1-year recurrent ischemic events. After adjusting for traditional risk factors, 1-hour GDF-15 change values greater than zero were associated with a 1.71-fold higher risk of outcomes compared to patients with change values less than or equal to zero (Table 4). Besides GDF-15 change values, only age (HR, 0.85; 95% CI, 0.77 to 0.95; *P*= 0.004) and BMI (HR, 1.02; 95% CI, 1.003 to 1.031; *P*= 0.01) emerged as independent predictors in the multiple model. Additionally, 3-hour GDF-15 change values showed a trend toward an increased risk of end points (HR, 2.45; 95% CI, 1.00 to 601.28; *P*= 0.05)(Table 4).

**Table 4.** Cox Regression Analyses for 1-Year Primary Events Rate in Relation to Biomarker Measurements.

| Measurements | Simple Model |  | Multiple Model <sup>a</sup> |  |
| --- | --- | --- | --- | --- |
|  | Hazard Ratio | P Value | Hazard Ratio | P Value |
|  | (95% CI) |  | (95% CI) |  |
| NT-proBNP with Evaluated Results, $\Delta > 0$ <sup>b</sup> | 2.28 (0.46-11.32) | 0.31 | 1.37 (0.21-8.90) | 0.74 |
| NT-proBNP with Evaluated Results, $\Delta > 0$ <sup>c</sup> | 2.50 (0.51-12.41) | 0.26 | 2.31 (0.20-26.30) | 0.50 |
| hs-cTnT with Evaluated Results, $\Delta > 0$ <sup>b</sup> | 2.62 (0.53-12.99) | 0.24 | 1.69 (0.27-10.45) | 0.57 |
| hs-cTnT with Evaluated Results, $\Delta > 0$ <sup>c</sup> | 4.35 (0.53-35.33) | 0.17 | 1.94 (0.30-12.69) | 0.49 |
| GDF-15 with Evaluated Results, $\Delta > 0$ <sup>b</sup> | 9.71 (1.19-78.91) | 0.03 | 1.71 (1.28-228.75) | 0.03 |
| GDF-15 with Evaluated Results, $\Delta > 0$ <sup>c</sup> | 2.21 (0.45-10.94) | 0.33 | 2.45 (1.00-601.28) | 0.05 |

HR, 95% CI, and *P* values were calculated with simple and multiple Cox regression analyses. <sup>a</sup> The
multiple models were adjusted for age, BMI, SBP, smoking status, history of hypertension, diabetes
change values categorized by zero. <sup>b</sup> The change values of GDF-15 between admission and 1 hour;
<sup>c</sup> The change values of GDF-15 between admission and 3 hours.

### 3.5 Incremental Prognostic Value of GDF-15 Temporal Evolution

The AUC of the clinical predictive model, including clinical risk factors and laboratory biomarkers, was 0.789. Adding GDF-15 levels on admission to the model did not result in a significant improvement in the AUC of the predictive model (AUC: 0.871, *P* = 0.365). When 1- and 3-hour GDF-15 change values were further added to the model, only the 1-hour GDF-15 change values improved the predictive ability of the model, with the AUC of 0.920 (*P* = 0.048) and 0.909 (*P* = 0.127), respectively (Fig 3).

**Fig 3.**
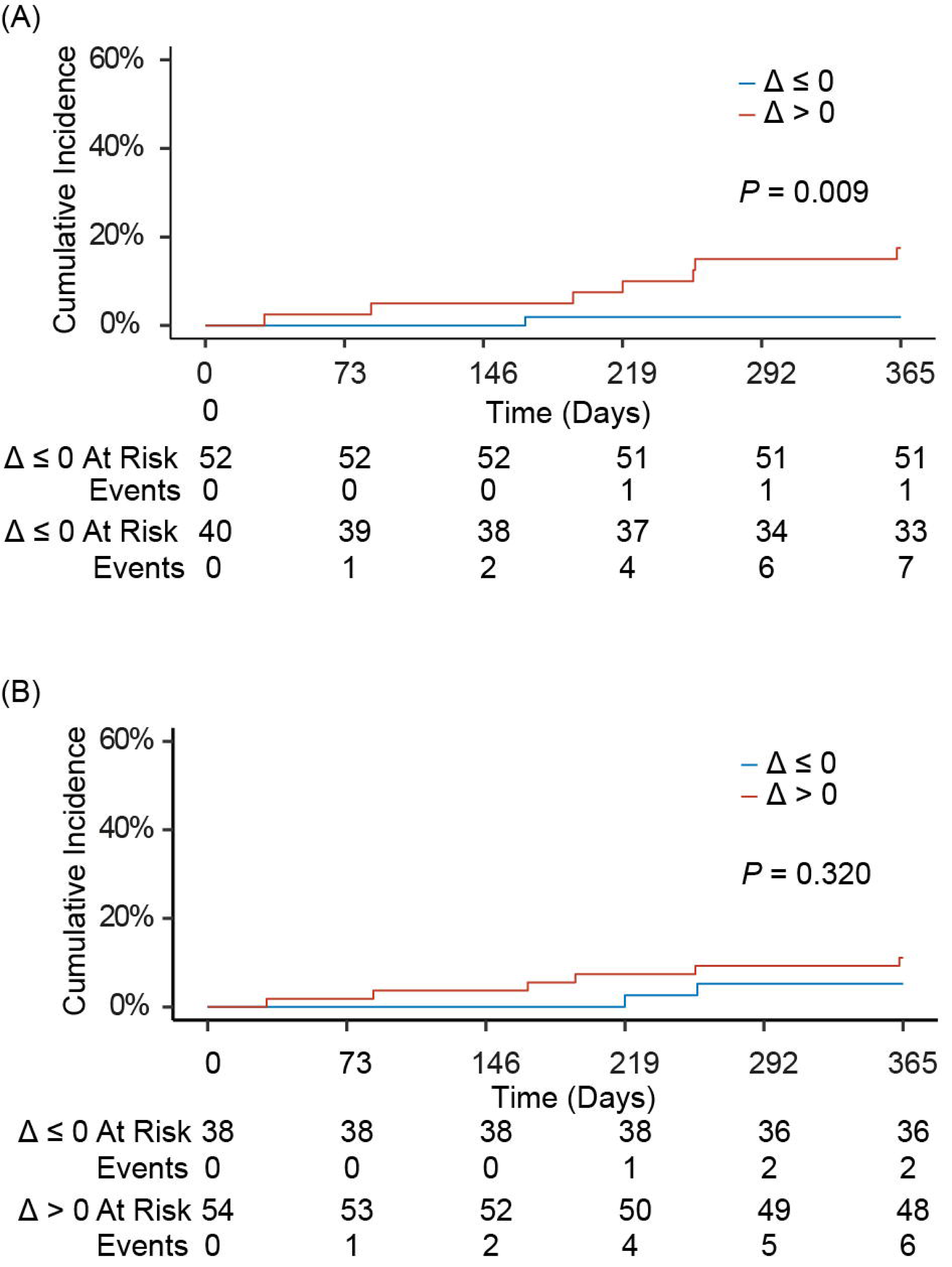
Receiver operating characteristic (ROC) curves for predictive models of recurrent ischemic events during 1-year follow-up. The x-axis represents specificity, and the y-axis represents sensitivity. Each colored line represents a different multivariable model. Model 1 (blue): adjusted for age, sex, BMI, SBP, smoking status, history of hypertension, diabetes mellitus, hyperlipidemia, myocardial infarction, chronic kidney disease and previous PCI; Model 2 (orange): Model 1 plus admission levels of hs-CRP, Cr, NT-proBNP and hs-cTnT; Model 3 (green): Model 2 plus baseline GDF-15 level at presentation; Model 4 (red): Model 3 plus 1-hour GDF-15 change (categorized as Δ ≤ 0 vs. Δ > 0); Model 5 (purple): Model 3 plus 3-hour GDF-15 change (categorized as Δ ≤ 0 vs. Δ > 0). These curves demonstrate the incremental predictive value of adding dynamic GDF-15 changes to clinical and biochemical risk models.

## Discussion

This study indicates for the first time that an increased level of GDF-15 1 hour during the acute phase, compared with the admission level, is an independent predictive factor for recurrent ischemic events within 1 year in patients with NSTE-ACS. Monitoring 1-hour changes in GDF-15 levels provides additional prognostic information beyond that offered by established clinical and biochemical markers.

Our study found that the median GDF-15 level in patients with NSTE-ACS was similar to that reported in previous studies [6, 11]. However, it was slightly lower compared to levels observed in patients with ST-segment elevation myocardial infarction (STEMI) [9]. Within 3 hours of admission, the circulating GDF-15 levels exhibited only minor changes compared to baseline, but a slight increase was observed. In STEMI patients, GDF-15 levels measured 90 minutes after the initiation of thrombolytic treatment increased significantly and were not associated with successful reperfusion [9]. Therefore, we hypothesize that the elevated GDF-15 levels in acute condition of ACS may reflect the extent of myocardial injury. Indeed, ischemia-simulated cardiomyocytes serve as a cellular source of circulating GDF-15 [4]. Moreover, a larger infarct size, as assessed by cardiovascular magnetic resonance, was associated with a major increase in GDF-15 levels following primary percutaneous coronary intervention in patients with STEMI [14]. This is further supported by our finding that the 1-hour change values of GDF-15 were correlated with the 1-hour change values of hs-cTnT.

GDF-15 levels on admission were independently related to cardiac dysfunction (NT-proBNP) and renal dysfunction (Cr), suggesting that GDF-15 concentrations may reflect cardiovascular risk by integrating the burden of inflammation, oxidative stress, and renal insufficiency. Unlike cardiac troponin, which is exclusively expressed in the heart [17], GDF-15 is not a cardiac-specific marker. For instance, GDF-15 is inducible in activated macrophages within human atherosclerotic plaques by oxidized low-density lipoproteins and their mediators [18]. Additionally, the R² value from the multiple linear regression was 0.31, indicating that these factors account for only about one-third of the variance in GDF-15 levels, while GDF-15 provides unique additional information. When GDF-15 was added to the prediction model consisting of traditional clinical risk factors and biomarkers, the AUC of the model increased.

Our study found that GDF-15 levels measured at admission and 1- and 3-hours post-admission were not predictive of the risk of recurrent ischemic events. While a single measurement of GDF-15 can independently predict the risk of mortality in patients with ACS, the conclusions of various studies regarding its ability to predict recurrent MI remain inconsistent [6, 9–14, 19–22]. Data from the GUSTO-IV trial showed that GDF-15 levels in patients with NSTE-ACS were not associated with the risk of reinfarction within 30 days after the index event, which the authors attributed to the short follow-up period5. Indeed, in another cohort with a 2-year follow-up, GDF-15 was identified as an independent predictor of the risk of recurrent MI [11]. However, the study populations in both studies were derived from large randomized controlled trials, and patient selection may have influenced the analysis of the predictive value of GDF-15 for recurrent MI. In an unselected cohort, GDF-15 was not found to be a reliable biomarker for predicting recurrent MI in patients with post-acute myocardial infarction [22]. Subsequently, the relationship between GDF-15 and specific types of recurrent events, including recurrent MI, is supported by a study with a large sample size and a well-characterized patient population [12]. The difference in findings among these studies may partly be explained by the different cohorts of patients in the studies, the timing of the plasma samples obtained, and the duration of follow-up. Our research demonstrated that increasing 1-hour GDF-15 levels were associated with an enhanced risk of recurrent ischemic events at 1 year. This suggests that serial measurements of GDF-15 may offer a complementary approach for predicting recurrent MI. However, it is noteworthy that the absolute difference change in GDF-15 levels from baseline to 6 months after stabilization of NSTE-ACS was not found to be associated with recurrent MI [13]. Combined with the conclusion of previous study, the importance of monitoring GDF-15 levels during the acute phase of the disease should be emphasized.

Some limitations should be mentioned. This is a single-center exploratory study with a small sample size, and its findings require validation in larger, more diverse patient populations to confirm the results and enhance the generalizability of the conclusions. Moreover, the sample size was insufficient to conduct separate analyses based on the type of treatment, due to the low number of events in each group. However, the study results indicated that there was no significant interaction between GDF-15 levels and intensive statin therapy in patients stabilized after ACS [12]. Although modeled on hs-cTnT 0/1h algorithms, the clinical utility of serial GDF-15 measurements, particularly within 1 hour, may be constrained by operational challenges in emergency settings. Moreover, our study did not include a healthy control group, as the design was specifically focused on prognostic evaluation within a diseased cohort. In addition, due to the lack of continuous monitoring of hs-CRP and Cr, as well as the stability of clinical risk factors such as age over a short period, we were unable to explore the relationship between the acute phase changes in GDF-15 levels and the changes and fluctuations in these variables.

## Conclusions

Based on the current results from our study, serial measurements of GDF-15 in the acute phase of NSTE-ACS serve as an independent risk predictor for recurrent ischemic events, providing incremental information beyond established risk factors. In line with the published literature [6, 10, 12, 13, 19, 20], we believe that GDF-15 is an important prognostic marker across all stages of coronary artery disease, from stable angina to acute instability and subsequent clinical stability. It is suggested that GDF-15 should be monitored throughout the course of NSTE-ACS to facilitate risk prediction. However, the timing and frequency of sample collection should be standardized to ensure consistency and reliability in its predictive value [23].

## Author Contributions

Ke Chen, Runan Zhang, and Hui Yuan: conception and design of the research. Ke Chen and Runan Zhang: acquisition of data. Ke Chen and Xinyi Rao: analysis and interpretation of the data. Ke Chen and Junyi Wu: statistical analysis. Hui Yuan: obtaining financing. Ke Chen: writing of the manuscript. Ke Chen, Runan Zhang, Xinyi Rao, Junyi Wu and Hui Yuan: critical revision of the manuscript for intellectual content. All authors read and approved the final draft.

## Data availability statement

The data that support the findings of this study are available from the corresponding author upon reasonable request.

## Acknowledgments

This work was supported by the National Key Research and Development Program (2022YFC2009600) (2022YFC2009602).

## Ethical standards

The study was conducted in accordance with the Declaration of Helsinki. The study was approved by the Ethics Committee of Beijing Anzhen Hospital, Capital Medical University (2021-072).

## Consent

For participants whose data could not be retrieved from electronic medical records, verbal consent was obtained via telephone, documented in writing and witnessed according to institutional guidelines. For participants whose anonymized data were available through the hospital system, the requirement for consent was waived by the Ethics Committee.

